# FGF21 defines a potential cardio-hepatic signaling circuit in human heart failure

**DOI:** 10.1101/2021.06.29.21259720

**Authors:** Salah Sommakia, Naredos H. Almaw, Sandra H. Lee, Dinesh K. A. Ramadurai, Iosef Taleb, Christos P. Kyriakopoulos, Chris J. Stubben, Jing Ling, Robert A. Campbell, Rami A. Alharethi, William T. Caine, Sutip Navankasattusas, Guillaume L. Hoareau, Anu E. Abraham, James C. Fang, Craig H. Selzman, Stavros G. Drakos, Dipayan Chaudhuri

## Abstract

**Background:** Extrinsic control of cardiac contractility and ultrastructure via neurohormonal signaling is well established, but how other organs regulate cardiomyocyte metabolism is less well understood. Fibroblast growth factor-21 (FGF21) a hormonal regulator of metabolism mainly produced in the liver and adipose tissue, is a prime candidate for such signaling.

**Methods:** To investigate this further, we examined blood and tissue obtained from human subjects with heart failure with reduced ejection fraction (HFrEF) at the time of left ventricular assist device (LVAD) implantation, and correlated serum FGF21 levels with cardiac gene expression, immunohistochemistry, and clinical parameters.

**Results:** Circulating FGF21 levels were substantially elevated in HFrEF, compared to healthy subjects (HFrEF: 834.4 ± 101.8 pg/mL, n = 40; controls: 145.9 ± 28.6 pg/mL, n = 20, p = 5.5 × 10^−8^). There was clear FGF21 staining in diseased cardiomyocytes, and circulating FGF21 levels negatively correlated with the expression of cardiac genes involved in ketone metabolism, consistent with cardiac FGF21 signaling. FGF21 gene expression was low in failing and non-failing hearts, suggesting at least partial extracardiac production of the circulating hormone. Circulating FGF21 levels were correlated with BNP and total bilirubin, markers of chronic cardiac and hepatic congestion.

**Conclusions:** Circulating FGF21 levels are elevated in HFrEF. The liver is likely the main extracardiac source, and congestive hepatopathy, common in HFrEF, was likely the proximate signal leading to FGF21 elevations. This supports a model of venous congestion from cardiomyopathy driving hepatic FGF21 communication to diseased cardiomyocytes, defining a potential cardio-hepatic signaling circuit in human heart failure.

## INTRODUCTION

A central feature of heart failure across different etiologies is a profound alteration in cardiomyocyte metabolism^1^. In heart failure with reduced ejection fraction (HFrEF), notable changes include increased reliance on glucose, ketones, and short-chain fatty acids, reduced pyruvate uptake by mitochondria and a consequent shunting of glycolysis towards the pentose-phosphate pathway^2-5^. Such adaptations are associated with altered expression of cardiomyocyte genes involved in the transport and metabolism of these different substrates. Some of these changes are dependent on cardiomyocyte-intrinsic signaling pathways triggered by cardiac dysfunction, but extrinsic control of cardiomyocyte metabolism is not well understood. Since extrinsic control via β-adrenergic and renin-angiotensin neurohormonal regulation is well established for cardiomyocyte contractility and ultrastructure, it seems likely such mechanisms exist to alter cardiomyocyte metabolism as well.

To explore extrinsic cardiometabolic signaling, we focused on fibroblast growth factor-21 (FGF21). This cytokine of the FGF19/21/23 family is produced primarily in the liver and adipose tissue, and is a potent regulator of fuel utilization and metabolism. Due to the absence of heparin-binding domains, secreted FGF21 can travel into the bloodstream and act as a hormone, binding to a receptor complex composed of a tyrosine kinase FGF receptor isoform and the β-Klotho (KLB) co-receptor. FGF21 regulates fatty acid oxidation in the liver, insulin sensitivity, glucose metabolism in adipose cells, and ketone usage^6^. In humans, FGF21 has been explored as a metabolic biomarker. In healthy subjects, it is induced late during the adaptive response to starvation^7^, but also during short-term carbohydrate overfeeding^8^, alcohol consumption^9^, and cold-induced thermogenesis^8^. Increased hepatic and adipose secretion is widely noted in diabetes and obesity, and skeletal muscle expression is noted after exercise or in hyperinsulemic states^10-14^.

Cardiomyocyte FGF21 signaling appears to be protective in several animal models of heart disease. In mouse models of hypertensive heart disease, deletion of FGF21 led to faster adverse cardiac remodeling, whereas application of exogenous FGF21 prevented cardiomyocyte hypertrophy^15, 16^. The effect seemed partly mediated by the reduction of fibrotic and inflammatory markers, but also by preventing a maladaptive decrease in PGC1α, the transcriptional co-factor regulating cardiac energy metabolism. In mouse models of myocardial ischemia-reperfusion injury, FGF21 deletion was associated with a greater reduction in cardiac function, which could be rescued by administering FGF21^17^. In this setting, improved cardiac function was associated with reduced markers of cardiomyocyte apoptosis. However, in studies of high-fat diets, mice were either protected or impaired after FGF21 deletion^18-20^. Nevertheless, data from these several disease studies seem to converge on a model revealing direct FGF21 action on the heart.

However, FGF21 signaling in heart disease remains unresolved in humans. In particular, whereas FGF21 appears protective in most animal models of heart disease, elevated levels are poor prognostic indicators in humans. In two studies of diabetic patients with coronary disease, higher levels of FGF21 were predictive of poorer outcomes^21, 22^. In cardiomyopathies, FGF21 elevations predicted adverse events in both heart failure with reduced or preserved ejection fraction, though >40% of patients in both these studies had diabetes^23, 24^. Additionally, whether the elevated blood FGF21 is synthesized in the heart itself or is produced in other organs remains unresolved. In mouse studies, FGF21 appears to be synthesized in diseased or metabolically-altered hearts, but not healthy cardiomyocytes, though cardiac expression was not seen in other studies^15, 16, 18, 25, 26^. Human data is scant, with one study showing an increase in FGF21 transcripts and two showing FGF21 staining in cardiomyocytes in alcoholic cardiomyopathy or hypertensive heart disease, all from a single group^26-28^. Thus, the extent of FGF21 elevation during HFrEF, what signals trigger this elevation, and whether it is active in the heart, remain unresolved questions.

Here, to investigate cardiac FGF21 biology independent of its well-established elevation during diabetes, we took advantage of cardiac tissue collection during the implantation of left ventricular assist devices (LVAD) in non-diabetic patients with end-stage HFrEF. First, we find that circulating FGF21 levels are elevated in HFrEF, and appear to act specifically upon failing cardiac tissue. Second, the concentration of circulating FGF21 correlates with markers of congestive hepatopathy, suggesting that the liver injury is a major trigger for its extracardiac secretion. Thus, FGF21 appears to be a novel hormone affecting cardiac function during periods of venous hepatic congestion.

## RESULTS

We retrospectively analyzed serum samples, obtained at the time of left ventricular assist device (LVAD) implantation, in 40 patients with ischemic and nonischemic HFrEF. Because we specifically wanted to determine the effect of heart failure on FGF21 levels, none of these patients had diabetes, end-stage renal failure, viral hepatitis, or non-alcoholic fatty liver disease (NAFLD), conditions known to alter FGF21 levels^11, 29-31^. A description of patient characteristics is included in Table 1.

**Table 1.**
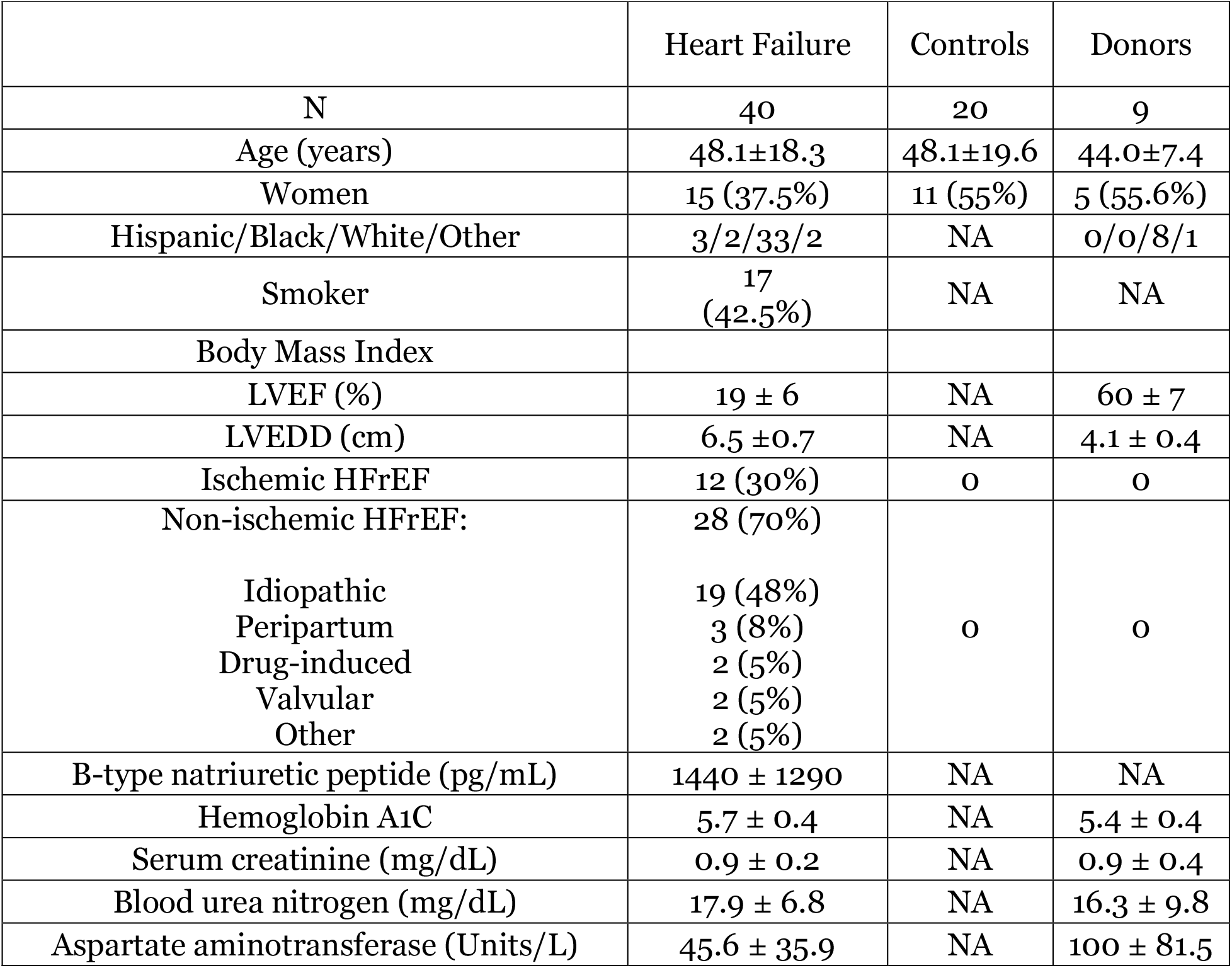
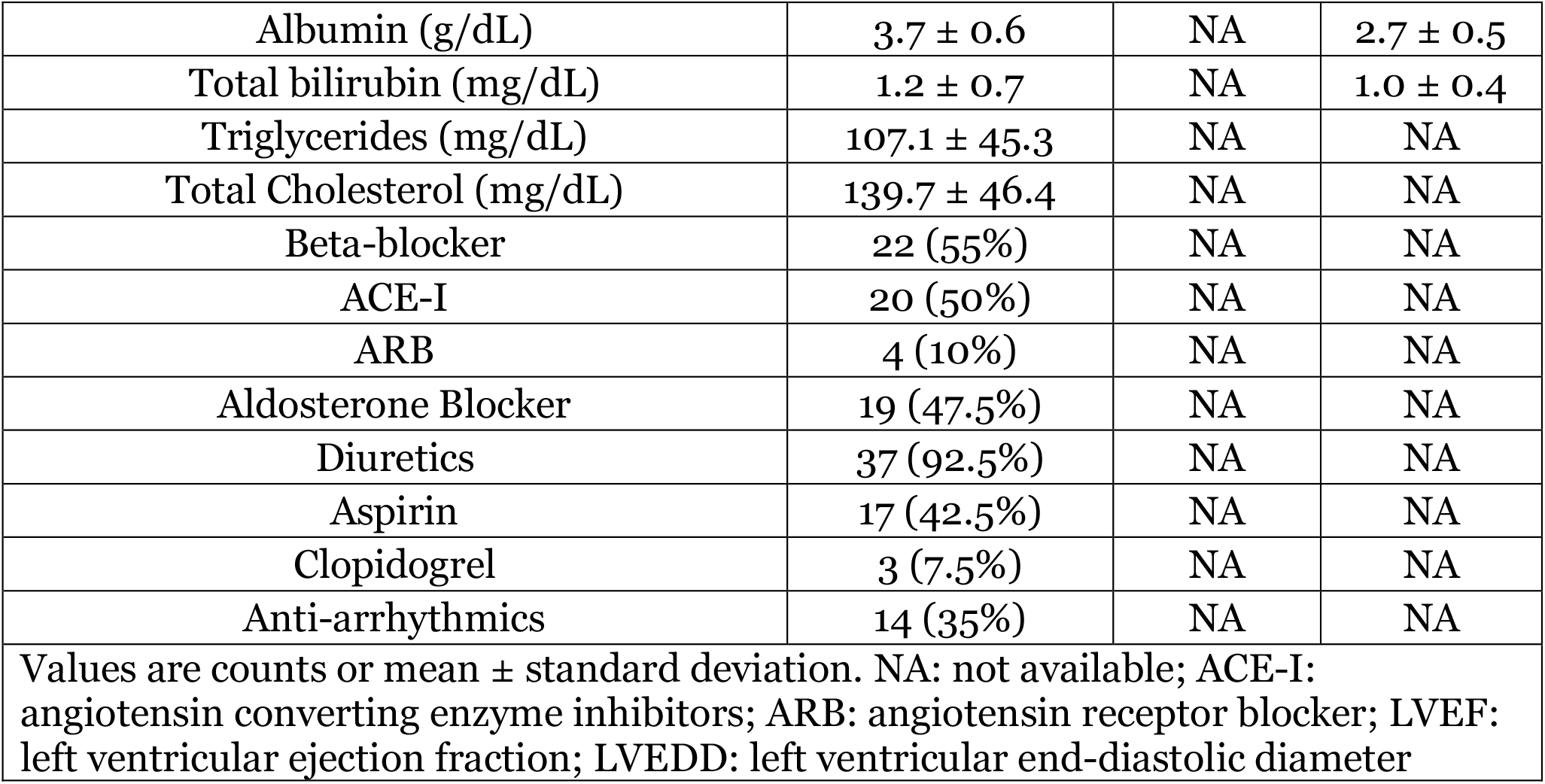
Characteristics of study population.

We measured blood FGF21 levels using an enzyme-linked immunosorbent assay. For controls, we utilized plasma from 20 age- and gender-matched healthy controls. Circulating FGF21 levels were more than fivefold higher in HFrEF patients compared to controls (834.4 ± 101.8 pg/mL vs. 145.9 ± 28.6 pg/mL, mean ± SEM, Figure 1A). In prior studies of FGF21 in human cardiomyopathies, a substantial fraction of patients had diabetes, a comorbidity expected to raise FGF21. Our results here show the increase in FGF21 during HFrEF is not due only to concurrent diabetes or NAFLD. Within the HFrEF group, no difference was observed in serum FGF21 between females and males (822.7 ± 156.5 pg/mL vs. 841.4 ± 135.5 pg/mL, Figure 1B), or between ischemic and non-ischemic etiologies (691.1 ± 133.9 pg/mL vs. 895.8 ± 133.4 pg/mL, Figure 1C). There was no correlation with the age of the subject (Figure 1D). In a subset of patients with mechanically-unloaded failing hearts, cardiac structure and function improves to the point that some of these patients can be weaned from mechanical support, and we assessed whether FGF21 level might predict such myocardial function improvement. In our cohort, recovery was defined as an improvement in left ventricular ejection fraction to >40% and reduction in left ventricular end-diastolic diameter to ≤59 mm. However, there was no significant difference in serum FGF21 between patients who recovered left ventricular function during mechanical unloading (responder, 774.2 ± 171.9 pg/mL, Figure 1E) and those that did not (non-responder, 860.2 ± 127.1 pg/mL).

**Figure 1.**
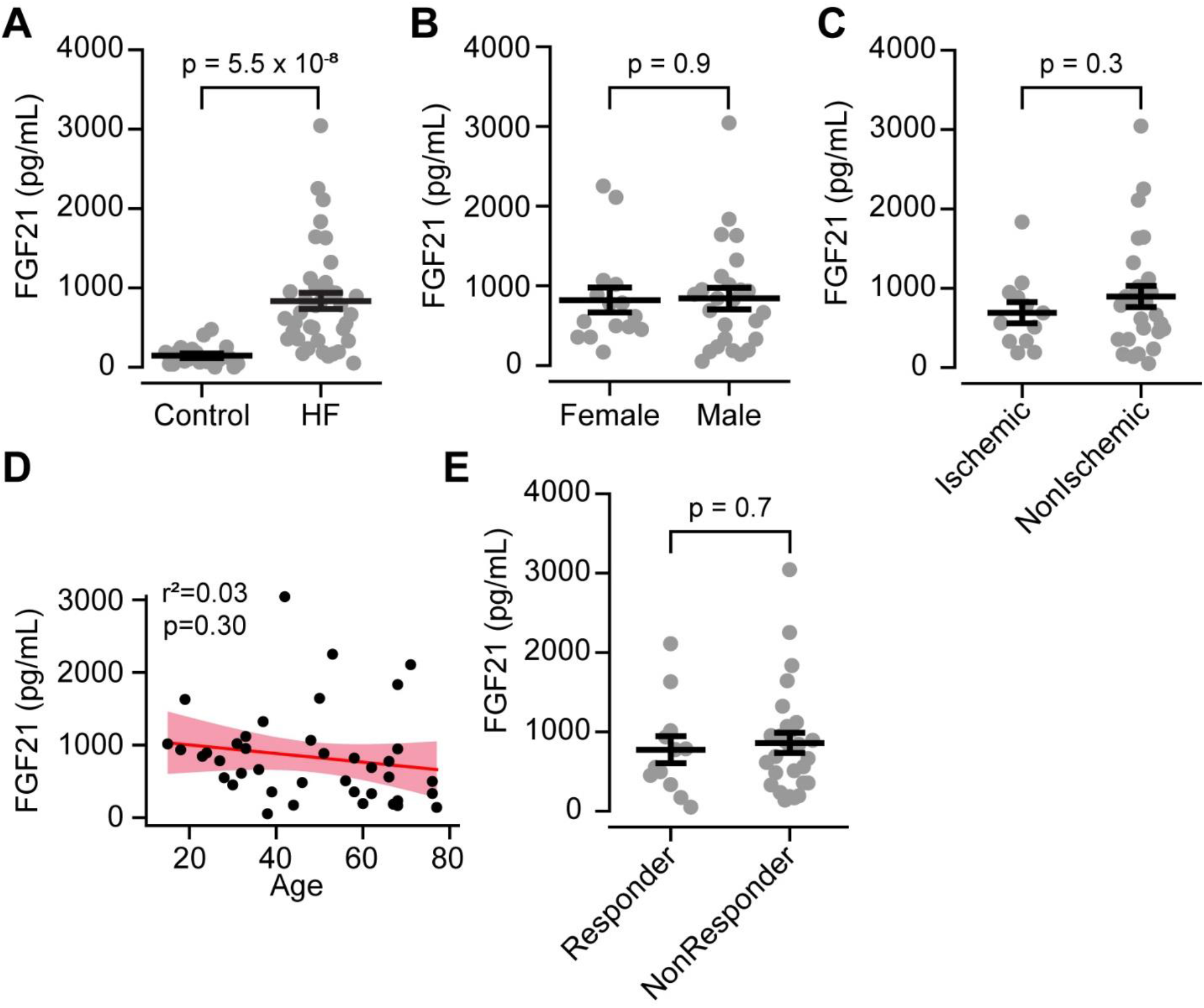
Circulating FGF21 levels are elevated in HFrEF. **A**. FGF21 levels in healthy controls (n=20) versus patients with HFrEF (n=40). **B**. No difference in FGF21 levels across gender in HFrEF patients (female = 15, male = 25). **C**. No difference in FGF21 levels between ischemic (n=12) and non-ischemic (n=28) HFrEF. **D**. No correlation with age of patient. Red line represents linear regression fit to the data, with pink shaded area representing 95% confidence bands. **E**. No difference in FGF21 between responders (n=18) and non-responders (n=22). Bars represent mean ± SEM. Each dot represents the level for an individual subject here and throughout.

In mouse studies of cardiac injury due to pathological hypertrophy, inflammation, and ischemia, FGF21 signaling has been seen within the heart^15, 26^, but whether such heart-specific expression occurs in humans remains unresolved, with a single study showing upregulation in cardiac tissue in people with end-stage heart failure^26^. We addressed two specific questions. First, whether FGF21 is found within heart tissue in HFrEF, which would define the human heart as a locus for FGF21 signaling. Second, whether the elevated serum levels are due to cardiac FGF21 synthesis.

To address the first question, whether FGF21 signals to the heart during HFrEF, we stained cardiac sections with anti-FGF21 antibodies in a subset of the HFrEF patients. As control, we examined cardiac sections in structurally intact hearts obtained from non-failing donors, but unused for human heart transplantation due to non-cardiac reasons. Although hearts in these donors were structurally and functionally normal, the donors themselves were deceased due to traumatic or anoxic brain injury, critical illnesses in which circulating FGF21 levels have been found to be elevated^32-35^. Our results replicated these findings, with donor serum FGF21 levels elevated (810 ± 225 pg/mL, n = 5) as in HFrEF serum. This set of conditions allows us to clearly identify if cardiac dysfunction leads to FGF21 signaling. In fact, whereas sections from donors showed essentially no cardiac FGF21 staining, sections obtained from HFrEF patients showed robust labeling throughout cardiomyocytes (Figure 2). This difference in cardiomyocyte staining is not due to altered levels of circulating FGF21, as serum levels were similarly elevated in both donor and HFrEF subjects. Rather, this result establishes that the failing heart is preferentially primed for FGF21-mediated metabolic signals.

**Figure 2.**
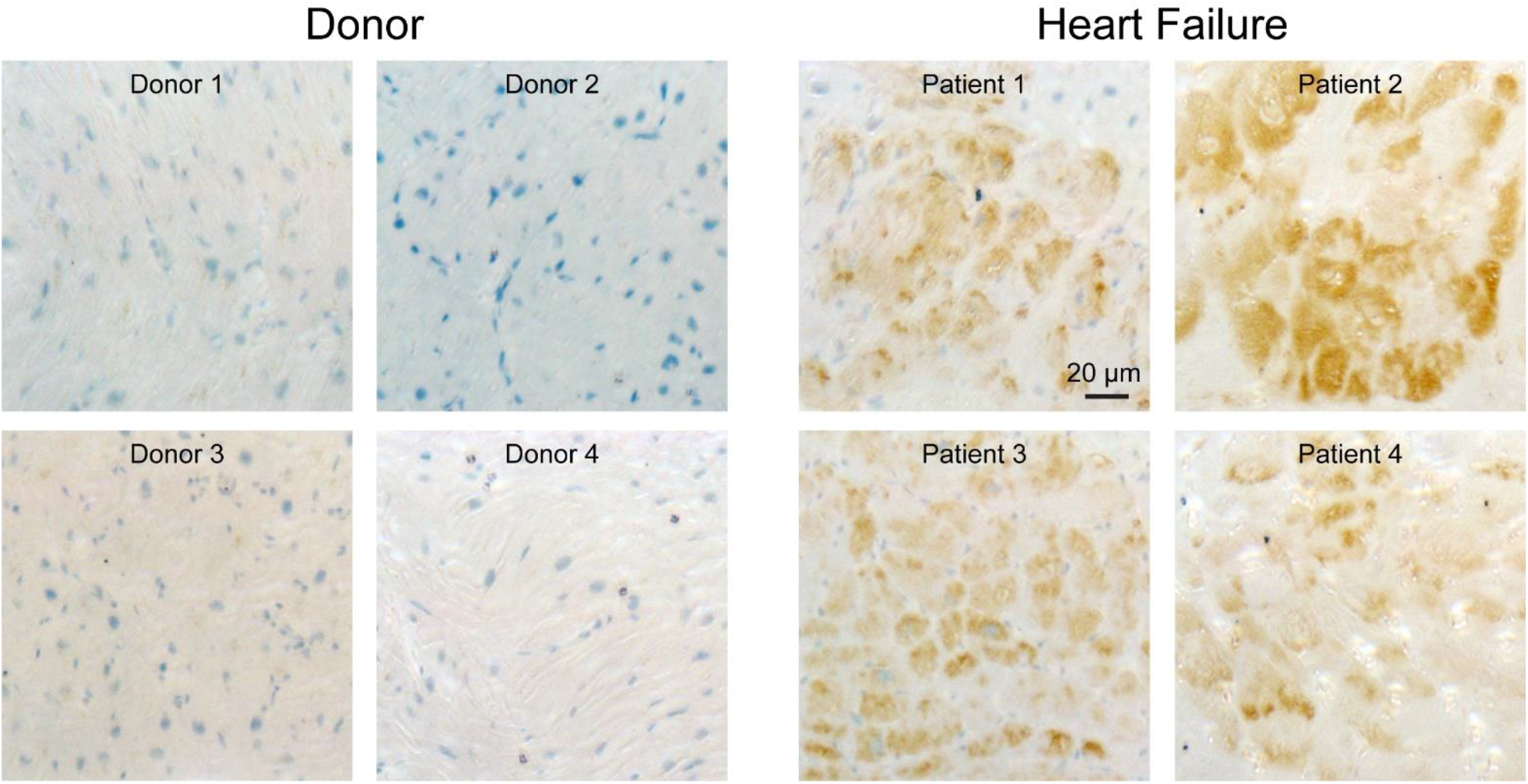
FGF21 is present in cardiomyocytes from HFrEF patients. Immunostaining for FGF21 in human heart slices reveals little FGF21 present in donor hearts but robust staining in HFrEF hearts. Nuclei are labelled in blue and FGF21 immunostaining is brown. Each panel is from a different patient.

Next, we addressed whether the FGF21 staining seen in heart sections represented a fraction bound from the elevated circulating levels or protein synthesized within cardiomyocytes. To address this issue, we assessed the expression of cardiac genes related to FGF21 signaling (Figure 3). Because the limited amounts of tissue obtained per patient are used in various assays across multiple studies, precluding Western blot analyses of protein levels, we restricted our analysis to measuring transcript levels via quantitative reverse-transcriptase polymerase chain reaction (qPCR). When we examined *FGF21* transcripts with qPCR, we found expression was near the limits of detection. Moreover, there was no clear change in expression between donor and HFrEF patients, nor any correlation with serum FGF21 levels (Figure 3), suggesting the *FGF21* gene may have low cardiac expression. To further assess cardiac *FGF21* transcription, we examined 7 published RNA-seq datasets obtained from cardiac tissue in human patients with ischemic, non-ischemic, restrictive, and hypertrophic cardiomyopathies (Table 2). *FGF21* transcripts were detectable in only 14 out of 167 samples, primarily in cardiomyopathy samples, and in most of these cases corresponded to 1-2 reads. Given limited cardiac FGF21 synthesis, it appears elevated circulating FGF21 during HFrEF may have a primarily extracardiac source. However, though FGF21 is synthesized elsewhere, it clearly signals to the heart, given the robust FGF21 staining we found in cardiac tissue in HFrEF, but not donors. Taken together, these results reveal an unexpected metabolic signaling axis to the heart from organs synthesizing FGF21.

**Figure 3.**
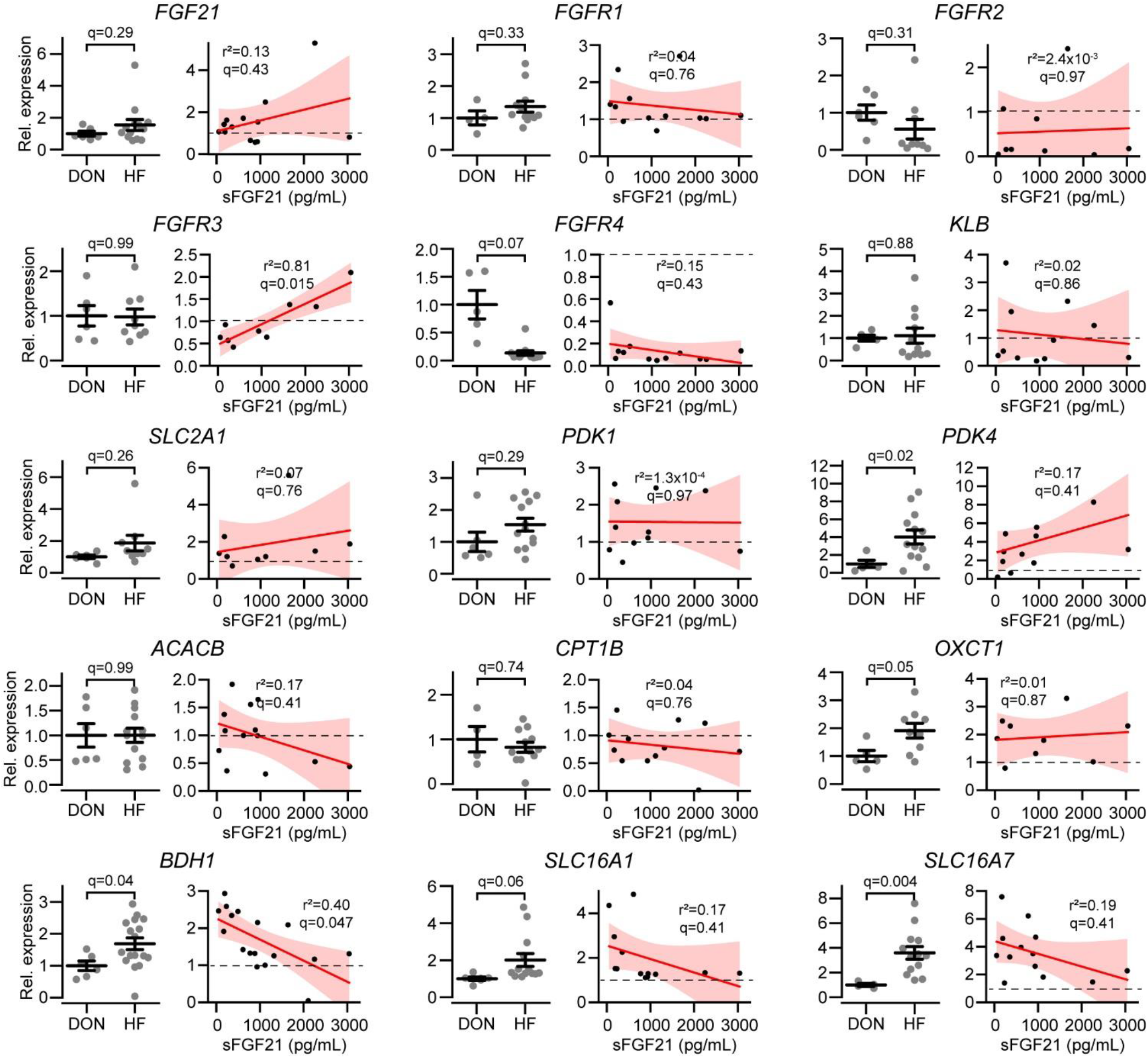
Correlations between serum FGF21 and cardiac gene expression. For each gene listed above, the left panel is the relative gene expression between normal donor (DON, n=4-6) and HFrEF hearts (HF, n=9-17). The right panel for each gene shows the linear regression for the HFrEF hearts against serum FGF21 (sFGF21) levels. The red line is the linear regression, with shaded area corresponding to 95% confidence bands. Coefficient of determination (r^2^) and corresponding Benjamini-Hochberg corrected p value (q) for a false discovery rate of 0.05 are shown.

**Table 2.** Human cardiomyopathy RNA-Seq datasets.

| Dataset (Ref.) | Cardiomyopathy Type | Total Samples | Detectable FGF21 transcripts – Non-failing (positive/total) | Detectable FGF21 transcripts – Cardiomyopathy (positive/total) |
| --- | --- | --- | --- | --- |
| GSE135055<sup>51</sup> | HFrEF | 30 | 0/7 | 0/23 |
| GSE71613<sup>52</sup> | HFrEF, Restrictive | 8 | 0/4 | 2/4 |
| GSE57344<sup>53</sup> | HFrEF | 6 | 1/3 | 1/3 |
| GSE46224<sup>54</sup> | HFrEF | 40 | 0/8 | 1/32 |
| GSE160997<sup>55</sup> | Hypertrophic | 23 | 0/5 | 2/18 |
| GSE133054<sup>56</sup> | HFrEF, Hypertrophic | 23 | 0/8 | 5/15 |
| GSE130036<sup>57</sup> | Hypertrophic | 37 | 0/9 | 2/28 |
|  |  | 167 | 1/44 | 13/123 |

As a first step towards examining cardiac FGF21 signaling in humans, we also assayed genes involved in the FGF response and fuel metabolism via qPCR (Figure 3). FGF21 exerts its effects primarily by binding to a receptor complex composed of one of four tyrosine kinase FGF receptor isoforms, typically FGFR1, and the β-Klotho (KLB) co-receptor^36^. In our hands, there was no evidence for a net increase in gene expression of FGF receptors nor the co-receptor β-Klotho between HFrEF versus donor samples. Intriguingly, however, there was a strong positive correlation between serum FGF21 and cardiac *FGFR3* expression. In contrast, most samples had suppressed *FGFR4* expression. We then turned to genes involved in the metabolism of glucose, fatty acids, and ketones. Compared to donor heart tissue, HFrEF cardiac samples had increased levels of *pyruvate dehydrogenase kinase 4* (*PDK4*), which inhibits the conversion of pyruvate into acetyl-CoA, as well as several genes involved in the transport and metabolism of ketones, including *solute carrier family 16 member 7* (*SLC16A7* or *MCT2*), a monocarboxylate transporter responsible for ketone uptake, *3-hydroxybutyrate dehydrogenase* (*BDH1*), which catalyzes the interconversion between the ketones β-hydroxybutyrate and acetoacetate, and *3-oxoacid-CoA transferase* (*OXCT1* or *SCOT*), which transfers the CoA group to the ketone acetoacetate. Unexpectedly, we found a negative correlation between serum FGF21 levels and *BDH1* transcripts, and similar but much weaker trends with the *SLC16A1* and *SLC16A7* transporters. This result was intriguing, as prior mouse studies revealed that inhibition of FGF21 reduced *Bdh1* transcripts, opposite to the correlation found here^37^. Although these analyses of correlations do not establish a particular mechanism for cardiac FGF21 activity, they do lend support to the hypothesis that FGF21 may act as a hormonal regulator of cardiac metabolism.

Our results established that serum FGF21 in HFrEF is elevated in the absence of overt metabolic diseases such as diabetes or NAFLD. To investigate the source of FGF21 production, we correlated serum FGF21 levels with the available clinical data on adiposity and cardiac, hepatic, and renal function (Figure 4). Notably, advanced HFrEF is associated with a cachectic phenotype, and elevated FGF21 levels have been attributed to muscle wasting during cardiac cachexia or prolonged fasting^7, 38^. In our cohort, however, we found no correlation with body-mass index (BMI) or aspartate aminotransferase (AST), a liver marker also released during muscle breakdown in cachectic states. Intriguingly, we observed the most significant correlation with B-type natriuretic peptide (BNP), followed by total bilirubin. Increased BNP is a sensitive marker for chronic pathological cardiomyocyte stretch, a condition associated with vascular congestion. Similarly, elevated total bilirubin in the absence of elevated liver function enzymes (AST) is the pattern encountered most frequently in congestive hepatopathy, which also occurs due to chronic vascular (venous) congestion. This suggests that chronic venous congestion may be a proximal signal for hepatic FGF21 secretion. Moreover, we observed a weak negative correlation with albumin, a marker of hepatic synthetic function, and stronger negative correlations with triglycerides and total cholesterol, which also partly reflect hepatic synthesis. We did not pursue correlations with invasive hemodynamic data, as these acute measures were optimized in the immediate preoperative period prior to LVAD implantation. Taken together, our data here supports the hypothesis that one main source of elevated circulating FGF21 observed in HFrEF is due to hepatic release.

**Figure 4.**
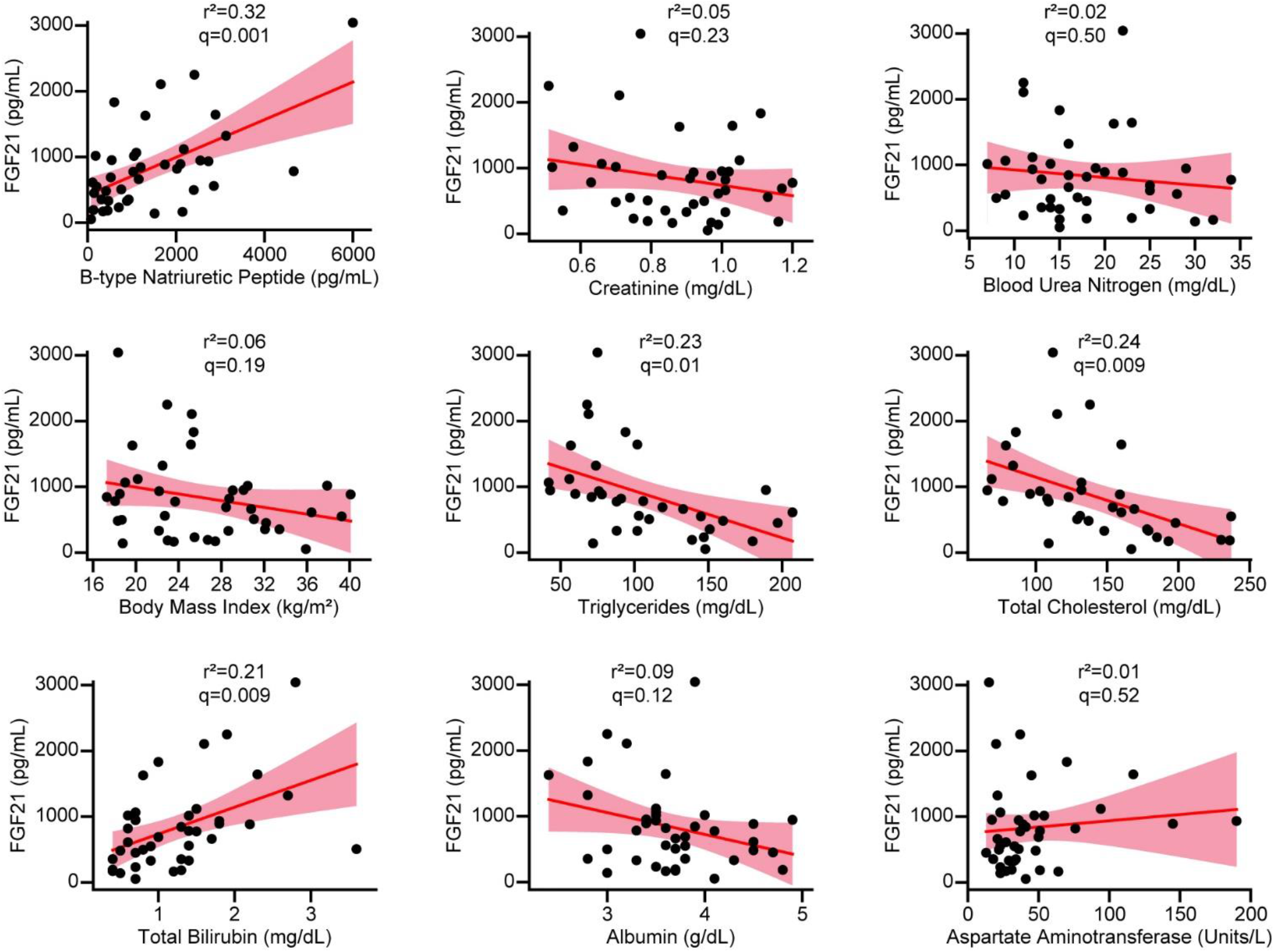
Correlations between serum FGF21 and clinical parameters. Correlation of serum FGF21 values with clinical index via linear regression (n=34-40). The red line is the linear regression, with shaded area corresponding to 95% confidence bands. Coefficient of determination (r^2^) and corresponding Benjamini-Hochberg corrected p value (q) for a false discovery rate of 0.05 are shown.

## DISCUSSION

This study describes three major findings defining human cardiac FGF21 biology. First, serum levels of FGF21 are elevated in the setting of HFrEF independent of other comorbidities, such as diabetes, that can raise hormone concentration. Second, FGF21 is present in cardiomyocytes from failing but not donor hearts, suggesting that it may activate downstream cardiac signals. Third, the origin of the elevated FGF21 appears to be partly extracardiac, with the liver as the most likely extracardiac source.

Elevated serum FGF21 levels have been seen in prior studies of heart failure with either reduced or preserved ejection fraction^23, 24, 38^. The aim of all three studies was to evaluate FGF21 as a biomarker for cardiac disease, rather than to elucidate human cardiac FGF21 biology. In two studies, diabetes was a significant comorbidity in >40% of subjects, confounding the ability to interpret the basis for elevated FGF21^23, 24^. In the third study, diabetic patients were excluded. In that study, serum FGF21 levels were elevated in 19 HFrEF patients and increased further in 19 subjects with HFrEF and cachexia, defined as a 5% non-edematous weight loss over 6 months. Notably, the HFrEF study by Shen *et. al*. was powered to detect cardiovascular outcomes, and elevated FGF21 levels predicted cardiac death independently of N-terminal pro-BNP levels^24^. More broadly, elevated FGF21 levels have been associated with adverse cardiac events in individuals with coronary disease and/or diabetes, though in subclinical disease such elevations are no longer predictive after adjustment for traditional risk factors^21, 22, 39, 40^. Thus, elevated FGF21 is a potential biomarker for severity in cardiac disease, raising the question of what derangements are causing this elevation, and whether the elevated FGF21 is an intrinsic or hormonal signal for the diseased heart.

The source of FGF21 signaling to the heart has been difficult to resolve. Initial studies suggested minimal expression in the mouse heart, whereas subsequent studies showed upregulation at both transcript and protein levels during cardiac hypertrophy^15, 41^. Conversely, in another study of FGF21 protection in a mouse model of hypertensive heart disease, circulating FGF21 was thought to arise from brown adipose tissue during adenosine receptor agonism^16^. In the only study of human cardiac gene expression, transcriptomic analysis revealed somewhat increased FGF21 expression during end-stage HFrEF^26^. In our qPCR data, whereas elevated transcription was seen in particular HFrEF samples, a uniform increase was not detected, corroborated by analysis of previously published transcriptomic datasets. Taken together, this suggests that cardiac FGF21 gene expression is low at baseline, with perhaps a slight, heterogeneous increase in cardiomyopathies.

Nevertheless, our data here is consistent with mouse and human data showing direct activity of circulating FGF21 on the heart^15, 18, 27, 28^. Robust cardiac FGF21 staining was seen in all the HFrEF samples but none of the non-failing donors, suggesting that elevated circulating FGF21 binds to the failing heart. In terms of cardiac regulation, in hypertrophy and ischemic models, exogenous FGF21 reduced fibrosis, inflammation, apoptosis, and maladaptive changes in cardiac energy metabolism^15-17, 19^. In this report, we queried a range of genes involved in cardiac metabolism. Surprisingly, we found an overall negative correlation between FGF21 and genes involved in the synthesis and transport of ketones. Whereas some prior data has suggested a direct relationship between FGF21 and fasting-induced ketogenesis, in humans the ketogenic response seems to precede the release of FGF21, suggesting a more circuitous relationship^7, 37^. Since our data is entirely correlational, we can only speculate that FGF21 acting on the heart may be a compensatory signal to preserve energetic homeostasis during progressive contractile failure. Intriguingly, there must definitely be changes in the response or susceptibility to circulating FGF21 during cardiomyopathies, as we saw no significant FGF21 staining in functionally normal hearts collected from donors, despite these individuals having similarly elevated serum FGF21 levels. In examining the FGF receptor family, we saw no obvious transcriptional upregulation that would indicate how FGF21 binds cardiomyocytes during HFrEF, though there was a strong positive correlation with *FGFR3*, and possibly transcriptional suppression of *FGFR4*.

Finally, we investigated the pathology responsible for elevating blood FGF21. The liver is the primary source of FGF21, though adipose tissue also produces it. It is likely the elevated FGF21 in HFrEF derives from the liver rather than adipose tissue, as we saw no relationship with BMI, and, when increased FGF21 levels are due to adipose release, it shows a positive correlation with lipid profile^11, 42^. Instead, here we find higher FGF21 associated with a lower lipid levels, which may reflect the triglyceride-lowering effect of exogenous FGF21^43, 44^. In regards to the mechanism activating hepatic FGF21 secretion, our data revealed a strong correlation between FGF21 and BNP levels, a marker of chronic myocardial stretch and vascular congestion. We also showed a correlation with elevated bilirubin but not AST, a pattern most consistent with congestive hepatopathy, which is also due to chronic vascular congestion. In summary, pathological hepatic venous congestion in HFrEF may be the proximate signal causing FGF21 release, with elevated FGF21 feeding back on the heart to regulate its metabolism. Thus, FGF21 defines a potential cardio-hepatic metabolic signaling loop in HFrEF.

## LIMITATIONS

First, although we assay a variety of clinical indexes, cardiac gene expression, and cardiac FGF21 protein, our analysis is based on correlations between these parameters, and a causal pathway has not been established. Without corresponding liver samples from these patients, we cannot confirm the serum FGF21 source is hepatic. Second, we did not study clinical outcomes. Our study was not powered or designed for clinical outcomes, as HFrEF patients went on to LVAD implantation and, for a substantial number of individuals, cardiac transplants. Finally, we studied a subset of patients with advanced HFrEF without diabetes, end-stage renal failure, or several other forms of liver disease. The biological activity of FGF21 may be more complex when these comorbidities are present, or when the early stages of heart failure are investigated.

## METHODS

### Study Population

The HFrEF cohort was taken from a larger sample of patients enrolled at the time of LVAD implantation. Patients who required LVAD support due to acute heart failure (acute myocardial infarction, acute myocarditis, and others) were prospectively excluded. Patients (age ≥ 18-years) were consecutively enrolled in institutions comprising the Utah Transplantation Affiliated Hospitals (U.T.A.H.) Cardiac Transplant Program (University of Utah Health Science Center, Intermountain Medical Center, and the Veterans Administration Salt Lake City Health Care System) with clinical characteristics consistent with dilated cardiomyopathy and chronic advanced heart failure who required circulatory support with continuous-flow LVAD. Non-failing donor hearts, not allocated for heart transplantation due to non-cardiac reasons (size, infection, and others) were used as controls. The study was approved by the institutional review board of the participating institutions, and informed consent was provided by all patients.

In the current study, we selected patients from the overall study who had blood samples available and who did not have diabetes (clinical diagnosis or hemoglobin A1c < 6.5%), overt renal failure (creatinine < 1.2 mg/dL), a clinical diagnosis of non-alcoholic fatty liver disease or viral hepatitis.

Blood samples used as the control reference were from healthy subjects recruited from the University of Utah and the surrounding Mountain West states. All subjects provided written, informed consent and all study protocols were IRB approved. Age- and gender-matched healthy donors were medication free and without acute or chronic illnesses.

### Blood and Myocardial Tissue Acquisition

Myocardial tissue was prospectively collected from the LV apical core at the time of LVAD implantation and was frozen before storage at -80 °C. Control samples were acquired from hearts that were not transplanted due to non-cardiac reasons. Donor LV apical tissue was harvested and processed the same way as the failing hearts. For HFrEF and donor samples, blood was collected immediately prior to the beginning of the operation. For healthy control samples, blood was collected by venipuncture. Samples were centrifuged and serum or plasma collected and stored for later analysis.

### Clinical Data Collection

Donor information like age, sex, and cause of death were collected with the help of DonorConnect. For HFrEF patients, clinical data including demographics, comorbidities, echocardiographic parameters, laboratory results, and other clinical data were collected within one week before LVAD implantation using our institutional research electronic data capture system (REDCap). We followed the echocardiographic data of each participant after surgery for up to 12 months. Based on left ventricular functional and structural changes following at least 3 months on LVAD support, patients were categorized as either responder or non-responder (see definitions in Results).

### Blood FGF21 measurements

Serum or plasma FGF21 levels were measured using the Quantikine ELISA Human FGF21 kit (R&D Systems, Minneapolis, coefficient of variation: Intra-Assay, 3.4%, Inter-assay 7.5%) according to the manufacturer’s instructions. Our control values were similar to prior studies conducted in either serum or plasma with this kit, and its accuracy has been validated in both^45-50^. Samples were run in duplicate. For each well, 50 μL standard or undiluted plasma or serum was added to 100 μL assay diluent, incubated for 2h at room temperature, washed 4X, then 200 μL conjugate was added to each well and incubated for 2h at room temperature. Wells were washed 4X, then 200 μL substrate solution was added and incubated for 30 min at room temperature, protected from light, then 50 μL stop solution was added to each well. Absorbance readings at 450 nm were recorded immediately following the addition of the stop solution, with a wavelength correction reading at 540 nm.

### Immunohistochemistry

The anti-FGF21 antibody (Abcam, ab171941) was used. Tissue pieces were fixed in 10% buffered formalin and embedded in paraffin. Sections 6 μm thick were made using a vibratome and mounted on coverglass. Sections were deparaffinized using Histosol (Fisher Scientific) then rehydrated by successively submerging in 100%, 95%, 70% ethanol followed by distilled H2O. Antigen retrieval was performed by submerging slides in antigen retrieval buffer (containing 60 mM sodium citrate and 40 mM citric acid, pH = 6.0) and heating in a microwave at a gentle boil for 10 minutes. Each section was then washed in wash buffer (containing 150mM NaCl, 50mM Tris pH=7.8, and 0.025% v/v Tween-20), permeabilized by incubation in permeabilization buffer (containing 150mM NaCl, 50mM Tris pH=7.8, 0.2% v/v Tween-20, and 1% w/v bovine serum albumin (BSA) for 15 minutes at room temperature, then washed 3X in wash buffer. Sections were then incubated in blocking buffer (containing 150 mMNaCl, 50 mM Tris pH=7.8, 0.025% v/v Tween-20, 10% v/v goat serum, and 1% w/v BSA) for 1 hour at room temperature, then in primary antibody buffer (containing 150 mMNaCl, 50 mM Tris pH=7.8, 0.025% v/v Tween-20, 1% w/v BSA, and 1:200 primary antibody) overnight at 4C. The following day, sections were washed 3X 5 minutes in wash buffer then incubated in oxidation buffer (containing 150 mMNaCl, 50 mM Tris pH=7.8, 0.025% v/v Tween-20, and 0.3%v/v H2O2) for 15 minutes at room temperature, followed by one wash in wash buffer. Sections were then incubated in secondary antibody buffer (containing 150 mMNaCl, 50 mM Tris pH=7.8, 0.025% v/v Tween-20, 1% w/v BSA, and 1:500 secondary antibody) for 1 hour at room temperature. Sections were then washed 3X with wash buffer and developed with DAB solution according to manufacturer’s instructions, washed 3X with wash buffer, then washed 3x with MilliQ H_2_O. Sections were counterstained with Mayer’s hematoxylin for 1 minute at room temperature, washed 3X with 10mM NaOH, then 3X with MilliQ H2O. Sections were then coverslipped using IHC mounting medium. Images were taken using an Olympus DSP camera mounted on an optical microscope.

### Cardiac FGF21 gene expression measurement

RNA was isolated from the LV tissue samples using the miRNeasy Mini Kit (Qiagen) or Purelink RNA Mini Kit (Thermo). Single strand complementary DNA (cDNA) was generated using Superscript Vilo IV master mix (Thermo) according to the manufacturer’s instructions using 1 μg RNA. cDNA was diluted 1:10 with Ultrapure distilled water (Thermo Fisher). Reactions were performed with 300 nM primers, using Power SYBR Green PCR Master Mix (Thermo) per the manufacturer’s recommendations. Quantification of gene expression was performed on a 96-well CFX Real-Time PCR System (BioRad). Analysis was performed by using the 2-ΔΔCt method, using the housekeeping gene *RPLP0*. Primers were based on prior publications, designed on NCBI Primer -BLAST, or obtained from Primerbank. A list of primers is below.

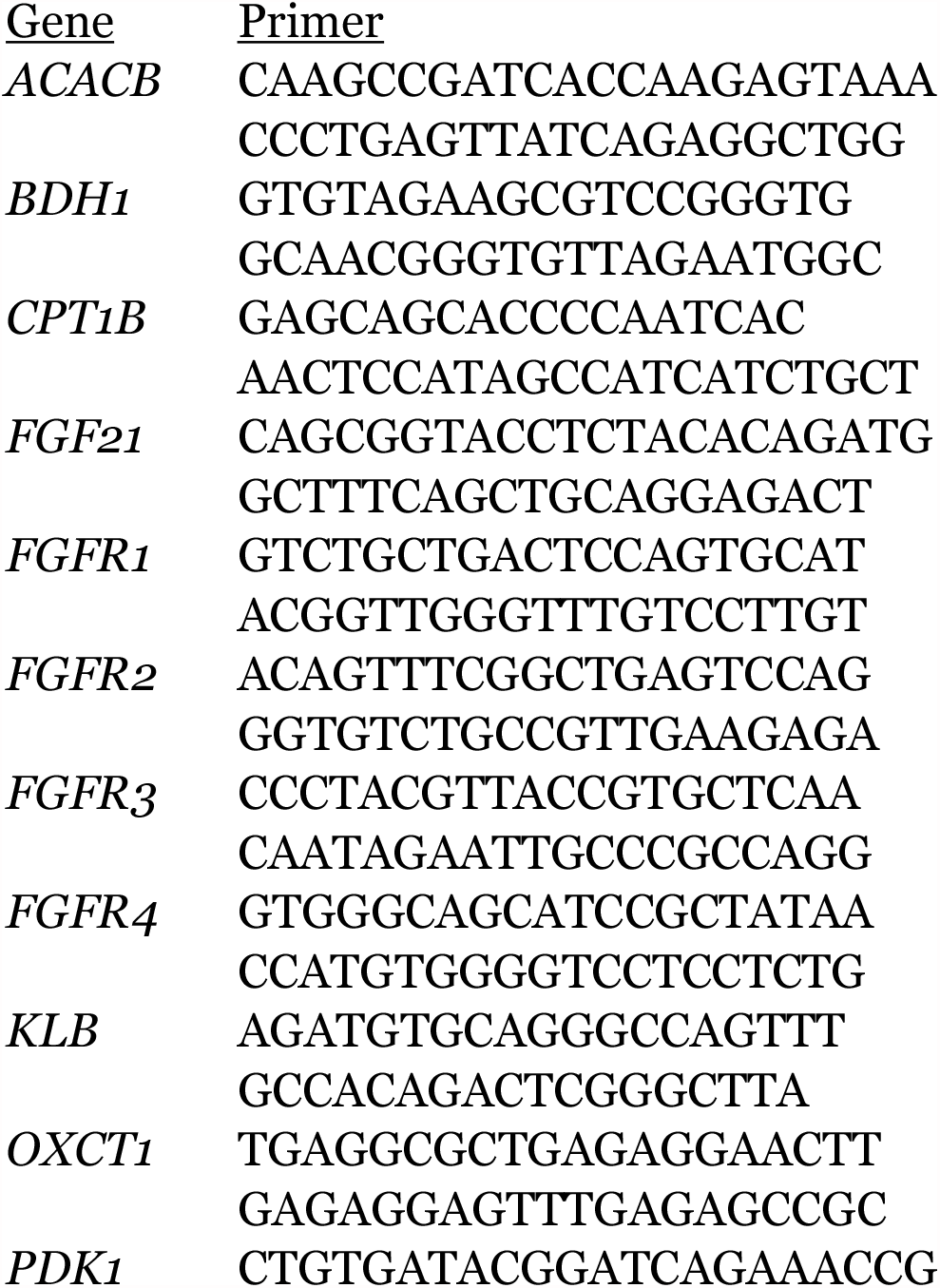
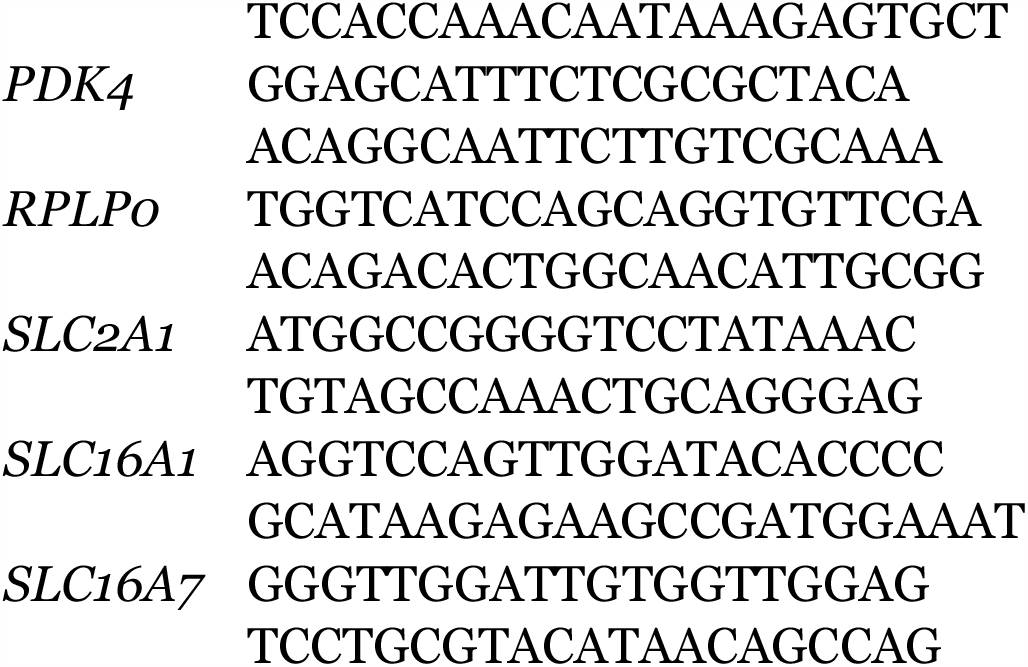

### Analysis of RNA-Seq Datasets

The NCBI Gene Expression Omnibus was searched for datasets obtained from human cardiac tissue, restricted to published studies on ischemic, nonischemic, hypertrophic, and restrictive cardiomyopathies using bulk RNA-Seq technology. We did not use single-cell RNA-Seq studies, because few were currently available and there is ongoing uncertainty about the interpretation of few or no reads per cell. We extracted metadata regarding samples, gene counting methods, and read format from the series matrix file, and raw or normalized count data for *FGF21* expression (searching for FGF21, ENSG00000105550, or ENST00000222157) from the supplementary data files. One dataset (GSE55296) was excluded as sample information was unavailable, one dataset (GSE65446) was excluded as the original study had been retracted. In these excluded studies FGF21 was either filtered out due to low expression or present in only the minority of individuals at low levels, similar to the presented data. The presented analysis includes seven datasets collected between 2014 and 2021.

### Statistics

Student’s *t*-test assuming unpaired, unequal variance samples were used for two-sample comparisons, assuming significance for p < 0.05. Correlations were performed using linear regression, with p-values for the coefficient of determination (r^2^) calculated from an analysis of variance using the *F*-statistic. For multiple hypothesis testing, a Benjamini-Hochberg procedure was used to derive corrected p-values (q-values) using a false discovery rate of 0.05. Analyses were performed in Excel and OriginPro 2020.

## Data Availability

Deidentified data will be available upon reasonable request.

